# The role of *Anopheles arabiensis* and *Anopheles funestus* in malaria transmission within houses with different characteristics in South-eastern Tanzania

**DOI:** 10.1101/2023.12.05.23299501

**Authors:** Alex J. Limwagu, Betwel J. Msugupakulya, Halfan S. Ngowo, Yohana A. Mwelugelo, Masoud S. Kilalangongono, Fredros O. Okumu, Bill E. Ngasala, Issa N. Lyimo

## Abstract

**Background:** Malaria vectors persist predominantly in houses lacking screened windows, doors, and eave space, leading to ongoing transmission. Some other vectors adapt their feeding behavior to sustain reproduction. This study evaluates the role of primary malaria vectors on malaria transmission in relation to house characteristics in Kilombero Valley.

**Methods:** Mosquito data collection was done using three methods: a CDC light trap, a miniaturized double net trap (DN-Mini trap), and a Prokopack aspirator through 222 households that were randomly selected in the three villages, namely Ebuyu, Chirombora, and Mzelezi. The houses are characterized during mosquito collections, and the Geographical Position System (GPS) was used to geolocate the sampled houses. Morphological identification was done on the collected sample (i.e., fed, part-fed, gravid, and unfed), and a sub-sample was sent to the laboratory to assess the sibling species, blood meal, and sporozoite.

**Results:** A total of 1542 Anopheline mosquitoes were collected, of which 79.1% (n= 1219) were *An. funestus* and 20.3% (n= 313) were *An. arabiensis* as primary malaria vectors, while the secondary malaria vector was 0.6% (n = 10). Out of 487 anopheline mosquitoes assayed for *plasmodium falciparum* sporozoite, 92.8% (n = 13) of *An. funestus* was reported to be dominant while *An. arabiensis* was dominant by 7.2% (n = 1). While *An. funestus* was the only species that had a blood meal and was found to feed on humans (88.16%, n = 134), dogs (11.18% n = 17), and a mixture of both human and chicken blood meal. In addition to that, the house with screened eave space had fewer *An. funestus* compared to the house with open eave space (RR = 0.978, p =0.864), while the house with a brick wall had a higher *An. funestus* compared with the house with a mud wall (RR = 0.690, p =0.107).

**Conclusion:** *Anopheles funestus* remained the dominant malaria vector responsible for all transmissions in the villages. The human population is at risk due to their preference for *An. funestus*, which contributes about 92.8% of infections in the village. Additionally, the house design appears to play a significant role in facilitating malaria vectors to continue transmitting malaria.

## Background

Malaria continues to be a major global public health concern, with an estimated 2 billion cases and 11.7 million deaths worldwide, particularly affecting Sub-Saharan African countries [1]. Malaria is primarily transmitted through the bites of infected female mosquitoes belonging to the *Anopheles* species. Four plasmodium species, namely *Plasmodium falciparum, Plasmodium malariae, Plasmodium vivax*, and *Plasmodium ovale*, are responsible for causing malaria [2]. *Plasmodium falciparum* was reported as the causative agent in 96% of malaria infections in Tanzania, particularly in malaria-endemic regions, while *Plasmodium malariae* and *Plasmodium ovale* accounted for the remaining 4% [3]. Tanzania has made commendable progress in reducing the malaria transmission rate, with prevalence dropping from 18.1% to 7.3% between 2007 and 2017 [3, 4]. However, during the COVID-19 pandemic, there was a slight increase of 1% in malaria cases due to healthcare system priorities and individual precautions [6]. Low- and middle-income countries, including Tanzania and Uganda, faced challenges in treating malaria during the pandemic [6, 7].

Furthermore, the persistence of malaria transmission in villages is influenced by various factors, including migratory farming. Families residing in temporary, semi-open shacks (shamba huts) for extended periods while working on distant farms are at higher risk of malaria due to inadequate mosquito protection [9]. Additionally, many houses in Kilombero Valley lack proper screening of eave spaces, windows, and doors, potentially allowing mosquitoes to enter homes [10–12]. Several studies suggest that modern house features are favored by residents due to the perception that these features reduce mosquito bites [13], However, in rural settings, concerns about the cost of maintaining these features have been raised. House improvement has been identified as an intervention that provides protection to all individuals in the house without involving insecticides [14]. In a study conducted by Bofu et al. in Kilombero in 2023, the community expressed the need for house improvement to protect families [13]. Despite reports of improved houses reducing malaria risk, there has been limited effort in national and international malaria strategies to prioritize housing improvement as part of malaria control.

The success in reducing malaria prevalence in Tanzania can be attributed to various interventions, such as the widespread use of long-lasting insecticide Nets (LLINs), indoor residual spraying (IRS), improvements in healthcare standards, and socio-economic enhancements, including better housing, education, income levels, and overall wealth [15]. Despite these efforts, residual malaria transmission persists in some rural areas of Tanzania [16]. One significant challenge contributing to malaria persistence is the insecticide resistance observed in *Anopheles funestus* mosquitoes, partly influenced by improper agricultural pesticide use [10, 11] Other factors, including malaria vectors, exhibit diverse behaviors that affect transmission[16]. Another important factor is the resting behavior of mosquitoes that are not resting on the targeted surfaces for indoor residual spraying [19].

In the 2000s, Kilombero and Ulanga districts primarily had *An. arabiensis* as the dominant malaria vector, despite *An. funestus* being more important [20]. Recent years have witnessed a significant shift in malaria transmission dynamics in Kilombero Valley, where *An. funestus* has become the predominant vector, responsible for over 85% of malaria transmission[21]. Both malaria vectors are developing resistance to commonly used chemicals [17,18,22].

Addressing malaria control in Kilombero Valley requires a multifaceted approach that targets *Anopheles funestus*, the current dominant primary vector, and addresses factors like healthcare access, housing conditions, and environmental management including appropriate applications of agricultural pesticide. This study aims to evaluate the role of malaria vectors in malaria transmission in regarding to house characteristics in Kilombero Valley.

## Material and Methods

### Study area

The study was carried out in Ulanga district, located in the south-eastern region of Tanzania (Fig 1). Mosquitoes were collected from three villages: Ebuyu (−8.979 S, 36.760 E), Chirombola (−8.926 S, 36.753 E) and Mzelezi (−8.898 S, 36.735 E). Average annual rainfall ranging between 1200 -1800mm and a mean annual temperature were 20–32 °C, as reported by Ngowo et al. in 2017 [23]. The primary economic activities in this area are rice and maize cultivation. Currently, Ulanga district faces the prevalence of two major malaria vectors, namely, *Anopheles arabiensis* and *Anopheles funestus*.

**Fig 1.**
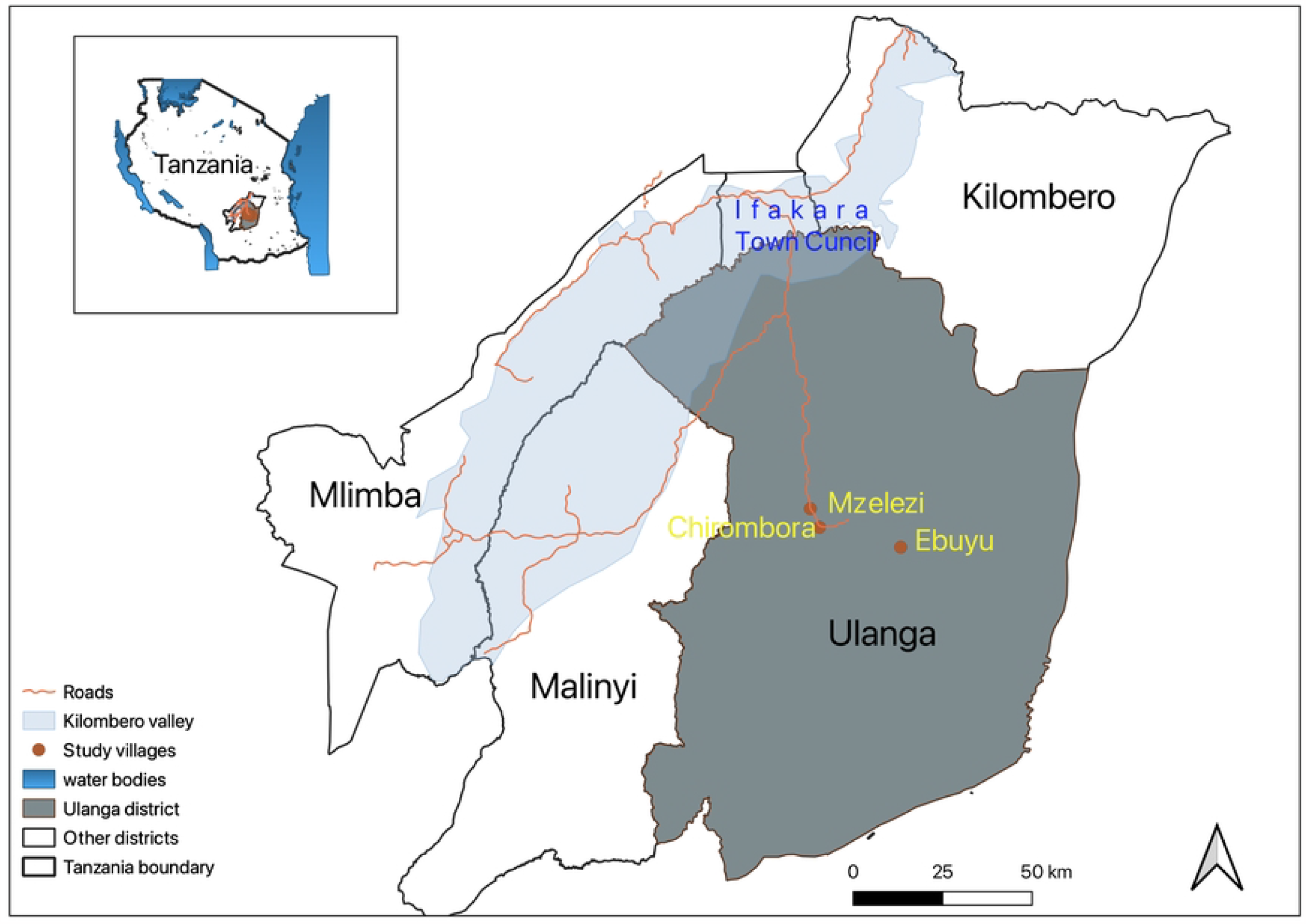
Map of the study area, showing study villages in Ulanga district, south-eastern Tanzania.

The typical housing structures in this region feature clay brick walls, open eaves (the space between the roof and walls), and open windows. *Plasmodium falciparum* stands as the predominant malaria parasite species in this area, while its transmission is primarily attributed to *Anopheles funestus* [21].

### Study design

#### Mosquito sampling

Adult anopheline mosquitoes were collected from March 2022 and July 2022. The host-seeking and resting mosquitoes were collected indoors and outdoors by using three methods: the double net trap (DN-mini trap) [24], CDC light trap [25], and Prokopack aspirator [26]. Mosquito data were collected for three days a week for four months. Here, the DN-Mini trap was used to sample mosquitoes indoors and outdoors, CDC-Light trap were used to collect host-seeking mosquitoes indoors, while a Prokopack was used to collect resting mosquitoes both indoors and outdoors. Mosquito collections were done from 6pm to 6am. The CDC light trap with a lid was hung about 1-1.5 m above the ground under occupied bed net. The DN-Mini trap was allocated in the sitting room, with volunteers inside acting as bait and collecting mosquitoes after every one-hour interval. Along with mosquito sampling, the sampled houses were also characterized, and variables such as eaves status, roof type, wall type, window status, presence of animals shed, latrine location inside or outside, and presence of poultry inside the house were collected from each sampled house. Geographical positioning system (Garming Extrex 20, GPS) coordinates were also collected during household characterization.

### Sibling Specie Identification of and Detection of Plasmodium Infection in Malaria Vectors

Female Anopheline mosquito collected were killed each morning using petroleum fumes, then sorted by taxa and physiological states [27], while the abdominal status of each female *Anopheles* mosquitoes was recorded as blood fed, unfed, partially fed or semi-gravid or gravid. Additionally, *An. arabiensis* and *An. funestus* were packed individually of one mosquito and in batches of ten mosquitoes in 1.5 ml microcentrifuge (Eppendorf®) tubes filled with silica gel as a preservative, then submitted to the laboratory for species identification by using multiplex polymerase chain reaction (PCR), sibling species of *An. gambiae s*.*l* [28] and *An. funestus* Koekemoer et al. [27]. and Cohuet *at al* [29]. Moreover, the head and thorax from *An. gambiae s*.*l* and *An. funestus* were separated from abdomen and tested for the presence of *Plasmodium falciparum* circum-sporozoite protein (*Pf* CSP) in their salivary glands, using the enzyme-linked immunosorbent assay (ELISA) method [27].

### House characterization

After collecting mosquitoes from traps placed both indoors and outdoors within the houses, participants recorded various house characteristics. These details included the house type, construction materials, and the overall condition of the house, encompassing factors like the presence of eave spaces, and the status of window and door screening. Additionally, precise house data was collected using a Global Positioning System (GPS) device and concurrently saved in a data collection form, which contained more information than what was initially stored in the GPS device.

### Blood meal analysis for Malaria vectors

Anopheline blood meal content analysis was conducted through an enzyme-linked immunosorbent assay (ELISA) using the abdomens of all blood-fed malaria vectors [30,31] to investigated for their blood meal sources, whether they were human, dog, chicken, or bovine. To prevent false positives, the ELISA lysate underwent heating to 100°C for 10 minutes, ensuring complete elimination of heat-labile non-*Plasmodium falciparum* antigens. Blood meal information from malaria-transmitting mosquitoes is crucial for understanding host preferences, transmission dynamics, and vector behavior.

### Data analysis

All data were analyzed using open-source statistical software, R program language version 4.2.11[32]. A generalized linear mixed model using template model builder (GlmmTMB) [32], was used to analyze mosquito count data. Here, we opted for GlmmTMB because of its ability to directly incorporate negative binomial and zero-inflated approaches. Mosquito counts were modeled as response variables, while house characteristics were modeled as fixed terms. On the other hand, household ID and sampling date were included in the model as random terms to account for any unexplained variations between houses and pseudo-replicates. Significance levels were considered at p<0.05.

### Ethical considerations

The research was conducted following the principles of the Declaration of Helsinki. Ethical approval was obtained from the Muhimbili University of Health and Allied Sciences Review Board (MUHAS-REC-12-2021-910), and permission to publish the work was granted by NIMR (Ref: NIMR/HQ/P.12VOL.XXXVI/35). Approval to conduct the study was obtained from the district medical officer of Ulanga district and the local government leadership in the selected villages. Before commencing the study, meetings were held with local government leaders to explain the study’s aim and procedures. Verbal and written informed consent were obtained from individual house occupants and human volunteers involved in mosquito collection. Participants were fully informed of potential benefits and risks, and their voluntary participation was assured. Respondents were informed of their right to withdraw from the study at any time without consequences, and confidentiality was maintained to ensure anonymity of participants

## Results

### Mosquito population and composition

Overall, a total of 19,841 female mosquitoes were collected indoors and outdoors between March and July 2022 using the CDC-light trap, DN-Mini trap, and Prokopack aspirator. *An. funestus* s.l. were 17.0% (n = 1219), *An. gambiae s*.*l*. were 1.6% (n = 313), and 0.1% (n = 10) were other anopheline mosquitoes. On the hands, *Culex* species were 99.8% (n = 18268) *Mansonia* were 0.2 %, (n = 29) and 0.0 % (n = 2) were *Aedes* species.

### Sporozoite Rate Infections of malaria vectors

Out of the 487 *Anopheles* mosquitoes submitted to the molecular laboratory for ELISA, 14 were confirmed to be infected with *Plasmodium falciparum* sporozoites. Among these infected mosquitoes, 92.8% (n = 13) were identified as *An. funestus s*.*s*., while only one mosquito, constituting 7.2% (n = 1), was classified as *An. arabiensis*.

### Effects of household characteristics on vector density

Regarding the mosquito collected in the house with different characteristics, the results show that, the house with metal roof material had fewer *An. funestus* mosquitoes with higher *An. arabiensis* compared to those constructed with grass roof material (RR= 0.646, p <0.05), (RR= 0.1600, p =0.5871), the house with ceilings had fewer number of *An. funestus* compared with those with ceilings (RR= 0.780, p =0.454), In addition to that, the house with screened eave space had fewer *An. funestus* compared to the house with open eave space (RR= 0.978, p=0.864) but not statistical significant, the house made by brick wall had higher *An. funestus* compared with the house made with mud wall (RR = 0.690, p =0.107) while the house with house with animals inside had fewer number of *An. funestus* compared to the houses no animals (RR= 0.672, p <0.01) Table 1.

**Table 1.** Effect of household characteristics on malaria vectors densities.

| Species | Variable (Household Characteristics) | Type | Mean ± 2SE | RR [95% CI] | p-values |
| --- | --- | --- | --- | --- | --- |
| *Anopheles funestus* | Roof Materials | Grass | 2.24 ± 0.972 | 1 | <0.05 |
| | | Metal | 2.10 ± 0.276 | 0.646 [0.418, 0.998] | |
| | Ceiling | No | 2.12 ± 0.272 | 1 | =0.454 |
| | | Yes | 2.00 ± 1.20 | 0.780 [0.407, 1.495] | |
|  | Eaves | Open | 2.02 ± 0.314 | 1 | =0.864 |
|  |  | Screened | 2.39 ± 0.498 | 0.978 [0.761, 1.258] |  |
|  | Wall Materials | Brick | 2.15 ± 0.285 | 1 | =0.107 |
|  |  | Mud | 1.76 ± 0.717 | 0.690 [0.439, 1.083] |  |
|  | Animal (100) | No | 2.47 ± 0.402 | 1 | <0.01 |
|  |  | Yes | 1.71 ± 0.331 | 0.672 [0.534, 0.847] |  |
| *Anopheles arabiensis* | Roof Materials | Grass | 0.206 ± 0.199 | 1 | =0.587 |
|  |  | Metal | 0.203 ± 0.083 | 1.600 [0.294, 8.715] |  |
|  | Ceiling | Open | 0.203 ± 0.079 | 1 | =0.575 |
|  |  | Screened | 0.2 ± 0.392 | 0.526 [0.055, 4.993] |  |
|  | Eaves | Open | 0.231 ± 0.096 | 1 | <0.01 |
|  |  | Screened | 0.120 ± 0.108 | 0.108 [0.022, 0.536] |  |
|  | Wall Materials | Brick | 0.206 ± 0.084 | 1 | =0.225 |
|  |  | Mud | 0.176 ± 0.154 | 0.351 [0.065, 1.899] |  |
|  | Animal (100) | No | 0.115 ± 0.080 | 1 | =0.254 |
|  |  | Yes | 0.301 ± 0.135 | 0.385 [0.074, 1.986] |  |

### Blood meal analysis

A total of 304 blood-fed *An. funestus* (identified by PCR) that were collected indoors and outdoors for both host-seeking and resting mosquitoes collection were tested for vertebrate host blood source (human, bovine, dog and chicken) from all three villages. Overall, the proportion of *An. funestus* that fed on humans was 88.16% (134/152), with dog blood being the most common non-human source Fig 2.

**Fig 2.**
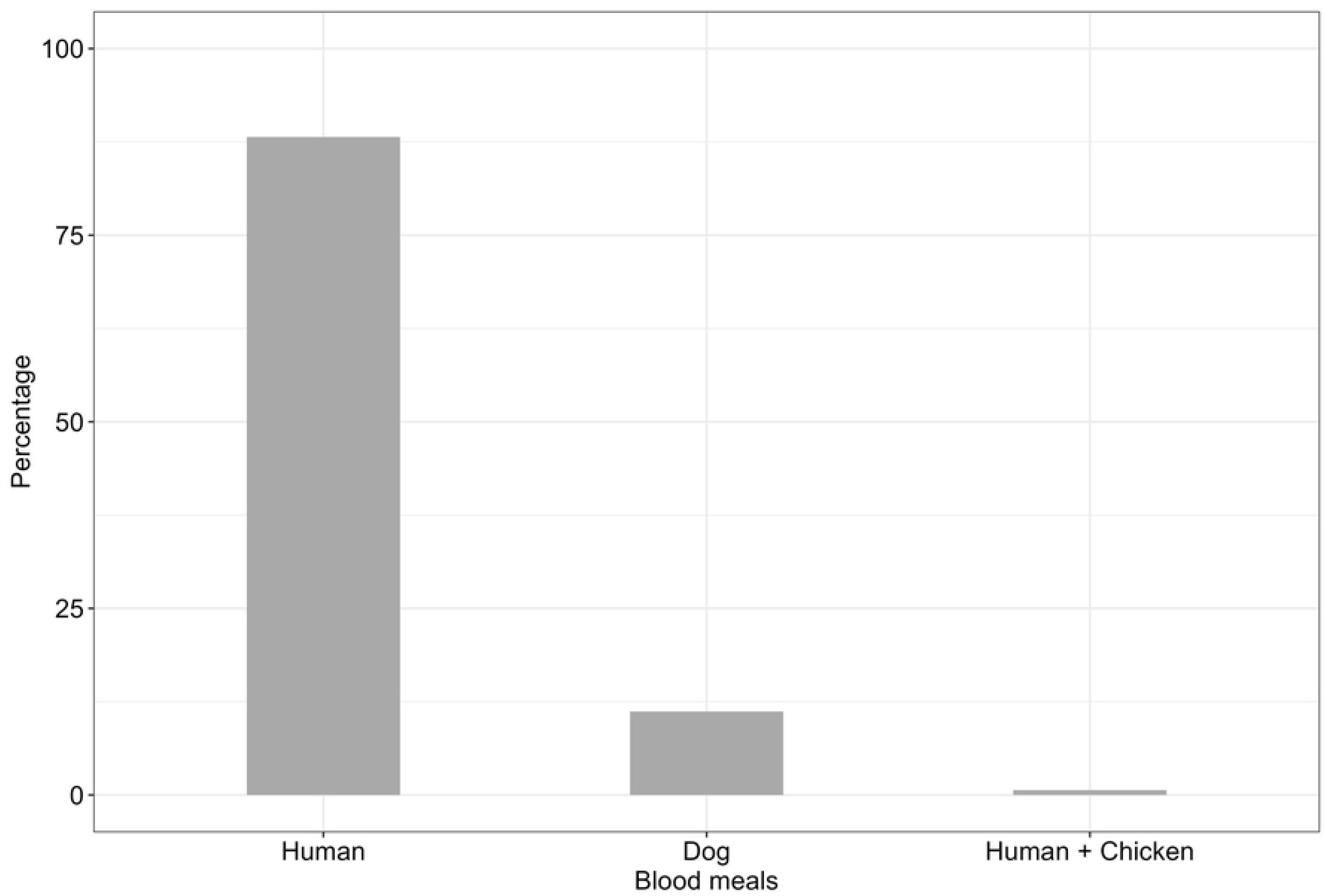
Showing a blood meal of *An. funestus* from different blood sources identified by ELISA method from the sample collected in Ulanga district, Mzelezi, Ebuyu and Chirombora villages.

### Ethical clearance and informed consent

The study was conducted following the principles of the Declaration of Helsinki. Ethical approval for conducting this study was obtained from the Muhimbili University of Health and Allied Sciences Review Board (MUHAS-REC-12-2021-910). while the permission to publish the work was approved by NIMR (Ref: NIMR/HQ/P.12VOL.XXXVI/27). Permission to conduct this study was secured through the approval of the District Medical Officer of Ulanga district and the cooperation of local government authorities in the chosen villages. Prior to commencing the study, comprehensive meetings were convened with local government leaders to elucidate the study’s objectives and methodologies.

Informed consent, both verbal and written, was diligently obtained from all individual residents of the households and the human volunteers engaged in mosquito collection. Participants were provided with detailed information regarding the potential benefits and risks associated with their participation, and their voluntary involvement was emphasized and respected. Participants were also assured of their right to withdraw from the study at any point without facing any adverse consequences, and strict measures were in place to maintain confidentiality, ensuring the anonymity of all participants.

## Discussion

This study shows that both *An. funestus* and *An. arabiensis* were the most prevalent mosquito species among the anopheline population in the study villages. Notably, *Anopheles funestus* predominated over to *An. arabiensis* in the rural Ulanga villages. However, a recent study revealed that, despite the lower population of *An. funestus* in comparison to *An. arabiensis* in the area, it accounted for over 85% of malaria transmission [21].

This study uncovered that *Anopheles funestus* mosquitoes not only carried the majority of malaria parasites but were also present in high numbers in all three villages. A previous study by Lwetoijera et al. in the Kilombero district also highlighted the increasing significance of *Anopheles funestus* in malaria transmission, particularly in the context of high resistance [33]. Furthermore, studies conducted in the Kilombero valley by Pinda et al, and Urio et al demonstrated that also *Anopheles arabiensis* also exhibited significant resistance to pyrethroid chemicals[18,22].

Based on our research findings, *Anopheles funestus* emerged as the predominant vector in terms of both malaria parasites and population density. In a prior study conducted by Kaindoa et al, [34], *Anopheles arabiensis* was identified as more abundant species. However, *Anopheles funestus* reported to harbor the majority of residual malaria parasites, even though it was in the minority compared to *Anopheles arabiensis* [34]. it seems that, *An. funestus* is increasing their dominance both in terms of abundance and transmission of malaria, which could be attributed to their resistance to insecticides used in malaria control.

Furthermore, our study revealed that *An. funestus* exhibited a strong preference for human blood meals over other potential hosts such as dogs and chickens. This anthropophilic behavior is consistent with the species known tendencies [35]. In recently study conducted in Kilombero valley in 2022, Katusi et al. revealed that, *An. arabiensis* and other secondary malaria vectors showed a preference for bovine blood over human blood. However, in another study conducted in the Kilombero valley, it was observed that *An. arabiensis* and *An. gambiae s*.*s*. exhibited a preference for feeding on chicken hosts over other available hosts [36]. Additionally, a separate study conducted in Kenya reported that mosquitoes deviated from the feeding on humans and opted for hosts other than humans, particularly in areas where livestock herding is prevalent [37], while other studies suggest that animal keeping may influence mosquito densities and potentially pose a risk to humans, while others argue that it could reduce malaria transmission risk [38].

In this study, we observed a relatively small number of *An. funestus* specie with the capability to feed on two different hosts as described in Table 1. where as *An. arabiensis* also have been commonly reported to have multiple feeding outdoor [16]. The tendence of mosquitoes feeding to various hosts has also been observed in neighbouring countries [39].

Our findings also indicate that dogs and chickens were the second most common sources of blood meals for *An. funestus* mosquitoes, following humans. This finding is in line with the prevalence of these animals in rural households, where dogs and chickens are commonly kept [35]. Studies by Katusi et al 2022 and Lyimo et al [35,36], reported that in Kilombero valley availability of cows influence the blood meal for *An. arabiensis* and *An. funestus*, like in Kenya found that, human blood were seems to be the most vulnerable to these species [40]. The preference for human blood meals by *An. funestus* is of particular concern for malaria control, as it increases the risk of malaria transmission to humans and underscores the importance of targeting this species in control strategies. Furthermore, it’s the observed that, higher *An. funestus* compared than *An. arabiensis*, suggest that *An. funestus* is the most responsible for malaria transmission in the area.

The study conducted by Lyimo et al 2013 [36], demonstrated that *An. arabiensis* was more likely to be affected by ivermectin, primarily applied to cattle, ivermectin affects the survival and egg production of *Anopheles* mosquitoes, consequently, this intervention appeared to have a controlling effect on *An. arabiensis* due to their behavior of feeding on other vertebrate blood including cows. On the other hand, there is limited knowledge regarding the impact of ivermectin on *An. funestus*, which exhibits a stronger preference for feeding on humans rather than cattle. *An. funestus*, being an indoor-biting species, is expected to be more susceptible to indoor interventions such as Long-Lasting Insecticide Nets (LLINs) and Indoor Residual Spraying (IRS), unfortunately, resistance to these interventions has been reported in this species.

This study also revealed that a substantial number of households had eave gaps and unscreened windows and doors Table 1, conditions that can facilitate mosquito entry and increase the risk of malaria transmission. These findings align with previous research which also emphasize the importance of addressing related factors in malaria control efforts [41].

Our study, also found that houses with eave space harbors more of *An. funestus* than those with no eave space, similar findings were reported by to Mburu et al 2018, where more Anopheline mosquitoes were caught in the houses with open eaves than other houses with small or single eave [14]. Many houses in the villages prefer to have eave space for air accelerations [42], but in other hands it poses a threat to malaria transmission risks, as mosquitoes are more likely to enter in the house through eave space. Several interventions on house screening have been introduced, this including the idea of eave tube treated with conventional insecticides plus house modification with eave tubes installed with physical barrier inside [42], treated curtains [43] and treated eave ribbons [35, 36], treated chair [46], treated sandals [47], all these tools introduced as structural interventions, in and around the houses, which may impact vector feeding, survivorship, population density and overall vectorial capacity and thus ability to transmit malaria and other pathogens.

This study has a limitation, where by a include small sample sizes was used from all three villages, therefore, further study is needed to study the major malaria vectors in the area, additionally, to investigate the factors that contribute to the rebounded of *An. gambiae* while they were eradicated by frontline interventions in Kilombero valley in 2000’s.

## Conclusion

*Anopheles funestus* is the prevalent malaria vector in the region and a significant 98% of this species were found to carry sporozoites. Additionaly, *Anopheles funestus* is one of the two species known to exhibit resistance to insecticides [17,18]. Several concurrent approaches [21,33,44,46–50], are required to complementary existing interventions like LLINs and IRS for effective malaria vector control specifically focus on targeting *An. funestus* specie.

House improvement and screening have also been contributing to malaria vector control in many developed countries [49]. Despite low quality evidence, the direction and consistence of effects indicate that housing is an important risk factor for malaria [51].

## Data Availability

All relevant data are within the manuscript and its Supporting Information file

## Acknowledgements

We extend our heartfelt appreciation to the village leaders and communities of Ulanga district for their invaluable cooperation and permission, which enabled the successful execution of this study. Our sincere gratitude goes to all the dedicated volunteers who played a crucial role in collecting the data. We offer special thanks to our project administrator, Rukiya Mohamed, for her exceptional support with logistical arrangements. We are deeply grateful to Marceline Finda for reviewing our manuscript, Najat Kahamba and Mohamed Jumanne for their valuable contributions during fieldwork. Additionally, we acknowledge the diligent efforts of Said Abbasi and Francis Tumbo in processing laboratory samples.

## Abbreviations

DN: Double Net
CDC: Centre for Disease Control
LLINs: Long Lasting Insecticides Nets
IRS: Indoor Residual Spray
GLMM: Generalized Linear mixed model
EMM: Estimated Marginal Means
ELISA: Enzyme linked immunosorbent assays
PCR: Polymerase Chain Reaction
GPS: Geographic Position System

## Availability of data and materials

Relevant data are within the paper. Also, data are available at: (DOI):

## Authors contributions

AJL was involved in DN-Mini trap design, study design, preparing the map of the study area, data collection, interpretations of the results, and drafting the manuscript. BJ was involved in the study design, data collection, and manuscript writing; MK was involved in the data collection; HN was involved in data analysis and manuscript review; FO was involved in the DN-Mini trap design, study design, supervision, and manuscript revision; BN was involved in the overall study design, supervision, and manuscript revision and IL was involved in the study design, supervision, and manuscript revision. All authors reviewed and approved the final manuscript.

